# Resting State fMRI for Motor Cortex Mapping in Children with Epilepsy

**DOI:** 10.1101/2022.02.04.22270184

**Authors:** Manu Krishnamurthy, Xiaozhen You, Leigh N. Sepeta, Emily Matuska, Chima Oluigbo, Madison M. Berl, William D. Gaillard, Taha Gholipour

## Abstract

Task-based fMRI is commonly used to localize motor functions prior to epilepsy surgery. Not all children are able to follow task instructions in the scanner, limiting fMRI motor mapping. We present a data-driven method to automatically delineate the motor cortex using task-free, resting state fMRI (rsfMRI) data of children with epilepsy and controls.

We preprocessed anatomical and rsfMRI images (64 patients, 12 controls) using fMRIPrep. We used FSL MELODIC tool for whole brain ICA estimation in multiple orders, to generate several sets of independent components. We defined a motor component selection process based on Discriminability Index-based Component Identification (DICI) score, resulting from comparing binarized component to the Human Motor Area Template (HMAT) in subject’s native brain space. Components with the largest DICI score and their peak in the template were combined to form the whole-brain ICA Motor Map (wIMM). We validated wIMM by comparing individual results with finger tapping motor task activation, and evaluated its reproducibility in controls with two runs of rsfMRI.

The hit rate between wIMM and motor task activation ranged 60 to 77% across all participants. Sensitivity of wIMM for capturing the task activation was 87.5% among 32 patients and 100% in 11 controls with available task results. The mean Dice similarity in results from repeated runs was 0.70 in controls.

Our results shows the sensitivity and reproducibility of an automated motor mapping based on ICA analysis of rsfMRI in children with epilepsy. The ICA maps may provide different, but useful, information than task fMRI. Future studies will expand our method to mapping other brain functions, and may lead to a surgical planning tool for patients who cannot perform task fMRI and help predict their postsurgical function.

## Introduction

Nearly thirty percent of epilepsy patients are considered drug resistant, and do not achieve adequate seizure control with anti-seizure medications alone (1). Surgery is the treatment of choice for many in this group. Identification of a seizure focus and its removal is associated with greater seizure control or freedom, and improved quality of life (2). The presurgical evaluation for epilepsy surgery aims to improve localization of the surgical target and minimize the risk of deficits in motor and language function from the surgery by mapping eloquent cortices. In recent years, task fMRI paradigms have decreased the reliance on and guided the invasive functional mapping methods using intracranial EEG or intraoperative electrical stimulation (3). For mapping hand motor cortex and its associated functional brain networks, finger-tapping task fMRI shows good correlation with invasive mapping using intracranial electrodes (4). Although task fMRI methods are feasible in most clinical circumstances, not all patients are able to follow task instructions in the scanner, even with an experienced functional imaging team. This can lead to spurious results encountered in evaluation of children or those with intellectual disabilities (5, 6). Resting state fMRI (rsfMRI) is acquired using similar imaging protocols as task fMRI but the patient is asked only to lay in the scanner and remain awake (7). Temporal correlation analysis of the rsfMRI signal can identify the normal resting state networks and largely overlap with networks elucidated through task fMRI (8, 9). These networks are consistent within and between individuals, across awake and non-awake states, and throughout development (10). With the advances in acquisition and analysis of fMRI, particularly network analysis through functional connectivity, rsfMRI is increasingly considered for developing clinical tools to complement or replace task fMRI for patients undergoing presurgical evaluation for brain surgery, including in epilepsy (6, 11, 12). The location of the target cortex can be displaced in patients with abnormal development such as childhood onset epilepsy. In order to capture atypical functional networks for clinical use, a data-driven method should demonstrate high sensitivity and specificity compared to conventional mapping. Independent Component Analysis (ICA) decomposes complex rsfMRI time-series data into distinct spatio-temporal components, without a seed region selection. Various studies have examined methods of identification of functional regions of interest using ICA and in different populations (13–16). However, ICA analysis lacks widespread clinical utilization in clinical practice. The development of clinical tools to map somatosensory cortex will provide the foundation for data-driven mapping of more complex brain functions. In this study, we implemented a whole-brain ICA method to map the motor cortex in children with typical development and those with epilepsy. We used an approach with automated identification of components of interest previously validated for language function in adults with brain tumors (14). In this study, to validate our approach, we compared ICA results to traditional task fMRI activation. We used motor mapping given the robust nature of activation and well-established neuroanatomy of motor functioning as a proof of concept, and hypothesize that our method will show good overlap with motor task fMRI results. To explore clinical utility, we also examined whether results correlated with available behavioral measures of motor functioning and seizure focus localization.

## Methods

### Imaging protocol

Images were collected with an 8-channel head coil on a 3T General Electric scanner. A 1mm isometric anatomical T1-weighted (T1w) image was collected during the same session for anatomical referencing and segmentation. For rsfMRI sequences, 60 participants were instructed to fixate on a cross on the screen with eyes open and avoid falling asleep, and 4 patients were instructed to watch a naturalistic video as visual stimulus (17). Each acquisition lasted six minutes for controls and five minutes for epilepsy patients. Images were collected using a gradient echo pulse sequence (3mm isotropic voxels, repetition time = 2s, flip angle = 90°, in-plane FOV = 192 × 192 mm). The motor task consisted of alternating 30s blocks of bilateral finger tapping and rest, serving as control in a blocked design. Motor task image acquisition parameters were the same as rsfMRI.

### Participants and Behavioral Data

Resting state fMRI from 12 children with typical development (“controls”, age 5-17) and 64 children with epilepsy (“patients”, age 10-23) from our clinical research database were retrospectively included in the study. Demographic and clinical characteristics are summarized in Table 1. A second rsfMRI run was available for the control group to evaluate reproducibility. In addition to rsfMRI, 11 controls and 32 patients had motor finger tapping task fMRI. As part of their pre-operative neuropsychological assessment, 64 patients had results from the 10-item Edinburgh Handedness Inventory (EHI) (18), 17 patients completed the Grooved Pegboard test, of which 15 patients completed the Delis-Kaplan Executive Function System (DKEFS) Trail-Making Test – Motor Speed condition (19). For those patients, motor hand dominance laterality index score derived from EHI, DKEFS scaled scores, and manual dexterity assessment standard score for each hand from Grooved Pegboard test were included. In EHI, Laterality Quotient (LQ) is calculated based on counting answers to each question as below:

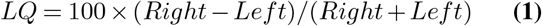

**Table 1.** Demographic and clinical characteristics of participants.

|  | Controls | Patients |
| --- | --- | --- |
| N | 12 | 64 |
| Sex (% female) | 41.7% | 51.5% |
| Handedness (% Right) | 83.3% | 60.9% |
| Left Hemisphere onset (%) | N/A | 54.7% |
| Right Hemisphere onset (%) | N/A | 42.2% |

A person is considered left-handed if LQ <-40, right-handed if LQ>40, and ambidextrous if |LQ|<=40. Studies were approved by Children’s National Hospital Institutional Review Boards and written informed consent was obtained from participants.

### Image Preprocessing

fMRI data preprocessing was done in fMRIprep 20.2.0 (20). Briefly, anatomical preprocessing included T1w bias field correction, brain extraction, and surface reconstruction. The T1w image was then volume-based nonlinearly spatially normalized to the MNI152Lin2009cAsym standard space. Functional images were coregistered to the T1w reference with six degrees of freedom, and head motion parameters were estimated before undergoing slice-time correction and resampling into native space and the MNI152NLin2009cAsym standard space. Motion outliers were defined as framewise displacement greater than 0.5mm or greater than 1.5 DVARS (20).For functional images, we excluded subjects with more than 1.3 minutes of motion outliers (3 controls and 9 patients).

### Whole brain ICA estimation

After smoothing rsfMRI data in participant’s native space using a Gaussian kernel (FWHM=6mm), we used the FSL Multivariate Exploratory Linear Optimized Decomposition into Independent Components (MELODIC) tool to estimate the ICA components (IC) (21). In brief, the 4D rsfMRI signal is decomposed into spatial and temporal components. The total number of components (TNC), which can be considered the order of the ICA decomposition, can be determined by the user, or recommended by the FSL melodic tool through a heuristic number recommended as “auto” or “default” number of components. The results are z-score probabilistic spatial IC (21). To account for individual variabilities in the integration of the resting state networks, we set six different TNC for ICA estimation: 20, 30, 40, 50, 60, and the “default’” value (Figure 1). We compared the results from all levels in subsequent steps to find the optimal TNC for each participant. We binarized each resulting IC at 11 thresholds of the ICs (z = 1-6, steps of 0.5) and compared (Figure 1).

**Fig. 1.**
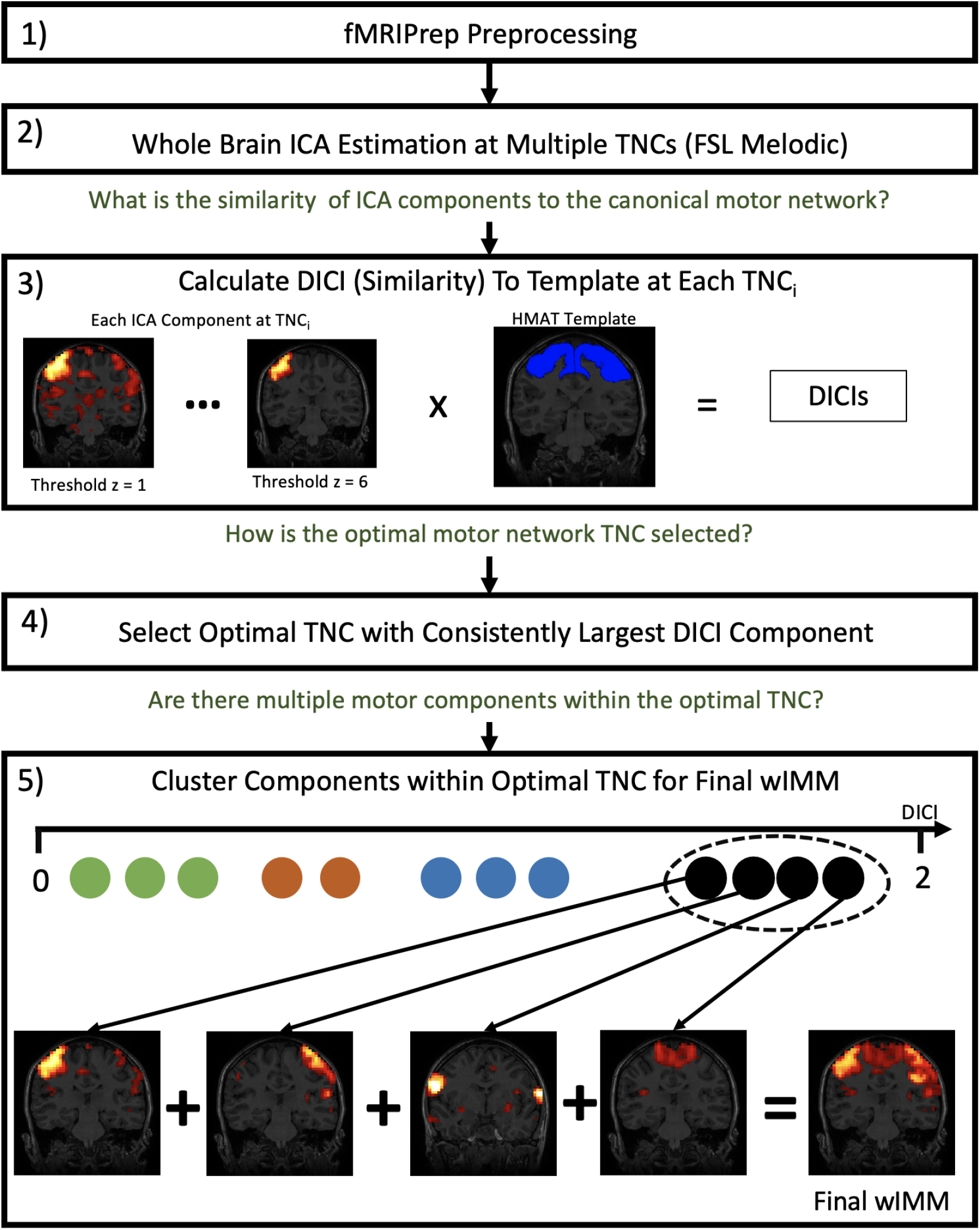
Overview of ICA motor mapping procedure for each subject. 1) Resting state fMRI image is preprocessed in fMRIprep. (2) MELODIC (FSL) ICA is used to generate multiple sets of components at a range of total number of component (TNC 20-60, default) parameters. (3) Within each TNC, components are binarized at different z thresholds (1-6) and the similarity between each binarized component and the HMAT motor area template in native space is measured by DICI. (4) The optimal TNC with the consistently largest DICI component across thresholds is selected for defining the final motor maps. (5) All components within the optimal TNC are clustered by their mean DICI value calculated across thresholds; the components in the largest DICI cluster with their peak in the template were combined to form the final wIMM.

### Template matching and component selection

To select the best components from the multitude of candidates from the prior step for each subject, we adapted an approach previously used in language network template matching method [Lu et al., 2017]. We chose Human Motor Area Template (HMAT) (22),a mask defining the most clinically relevant regions of the sensorimotor network. We transformed the HMAT template into the native space of each subject using Advanced Normalization Tools (ANTs). At each of the 6 different TNC levels, we compared generated components to the HMAT template using the Discriminability Index-based Component Identification (DICI) measure (14, 23):

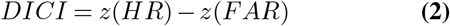

Where HR is hit rate and is calculated as the overlap between the component and template divided by the template region, measured in voxels. False alarm rate (FAR) is calculated by the number of the component’s voxels inside the brain but outside the template, divided by the total number of voxels outside of the template. The sensitivity of each component for matching the template is represented by HR and the specificity of the components is equal to 1-FAR, thus DICI includes both sensitivity and specificity, providing an objective measure to compare and identify the most likely component(s) matching the template. The z-scores of the hit rate and false alarm rate are calculated using an inverse Gaussian distribution. After DICI calculation for each IC, the TNC with the consistently largest DICI component across thresholds was chosen to define the final motor maps. To address possible splitting of the best motor component into multiple ICs, we used a mean shift clustering function (24) to cluster the mean DICI across thresholds of all components within the selected TNC. The components in the largest value cluster with their peak voxel contained in the template were combined to form the final resulting whole-brain ICA Motor Map (wIMM).

### Component validation by comparing to task activation

We compared the wIMM to the task-activated motor regions during the finger-tapping task. For task activation results, the preprocessed and spatially smoothed task fMRI data underwent single subject first-level activation analyses in SPM12 using GLM in native space with motor task vs control block set as contrast. The position of the peak task activation was recorded, and the resulting T-score map was thresholded to correspond to significance level of p<0.05 and constrained to the HMAT motor template and used as the task activation for comparison. We calculated the percent overlap between task activation and the wIMM (thresholded at z = 1.96) at the individual level, then calculated average overlap across subject groups. We also examined if individual wIMM always captured the task activation peak (i.e. sensitivity), and reported the percent of participants in each group with their peak task activation position included in their wIMM. To evaluate the effect of the seizure focus on the symmetry of the wIMM and task results, we calculated the laterality index (LI) for wIMM and task activation results after applying a threshold of z=1.96 (p<0.05), and compared the wIMM and task activation LI between patients with left- and right–onset seizures.

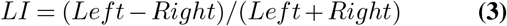

To understand the impact of hemisphere onset on results, we also compared the wIMM and task activation LI between patients with left- and right–onset seizures.

### Clinical validation

We compared wIMM and task activation results based on their correlation with available neuropsychological scores. Standardized motor function scores for the dominant and non-dominant hands based on the Grooved Pegboard neuropsychological measure, the scaled trail-making motor score from the DKEFS, and the EHI were included.

### Subject-level reproducibility

To evaluate the within-subject reproducibility and robustness of the whole brain ICA method, we repeated the pipeline for a second run of rsfMRI from the same session in the control group. We calculated the Dice coefficient of the wIMM (z threshold 1.96) across runs and compared left hemisphere, right hemisphere, and bilateral mean z value of the motor regions, and motor component map LI across runs.

### Statistical analysis

The wIMM pipeline was written and performed in MATLAB and all subsequent statistical analyses were conducted in R. Codes are available through https://github.com/tgholipour/ICAmotor.

## Results

### Individual whole brain ICA motor mapping

The averaged wIMM and the distribution of individual maxima are illustrated in Figure 2. We observed higher percentage of volumes meeting motion outliers definition in controls, compared to patients (t(df) = -2.31(12.07), p = 0.04). The majority of motor components resulted from ICA estimation with a TNC of 60 for patients and TNC of 20 components in controls. Neither TNC nor the number of components combined to generate the final wIMM were correlated with subject movement in the scanner defined as the percent of volumes designated as motion outliers (p>0.05).

**Fig. 2.**
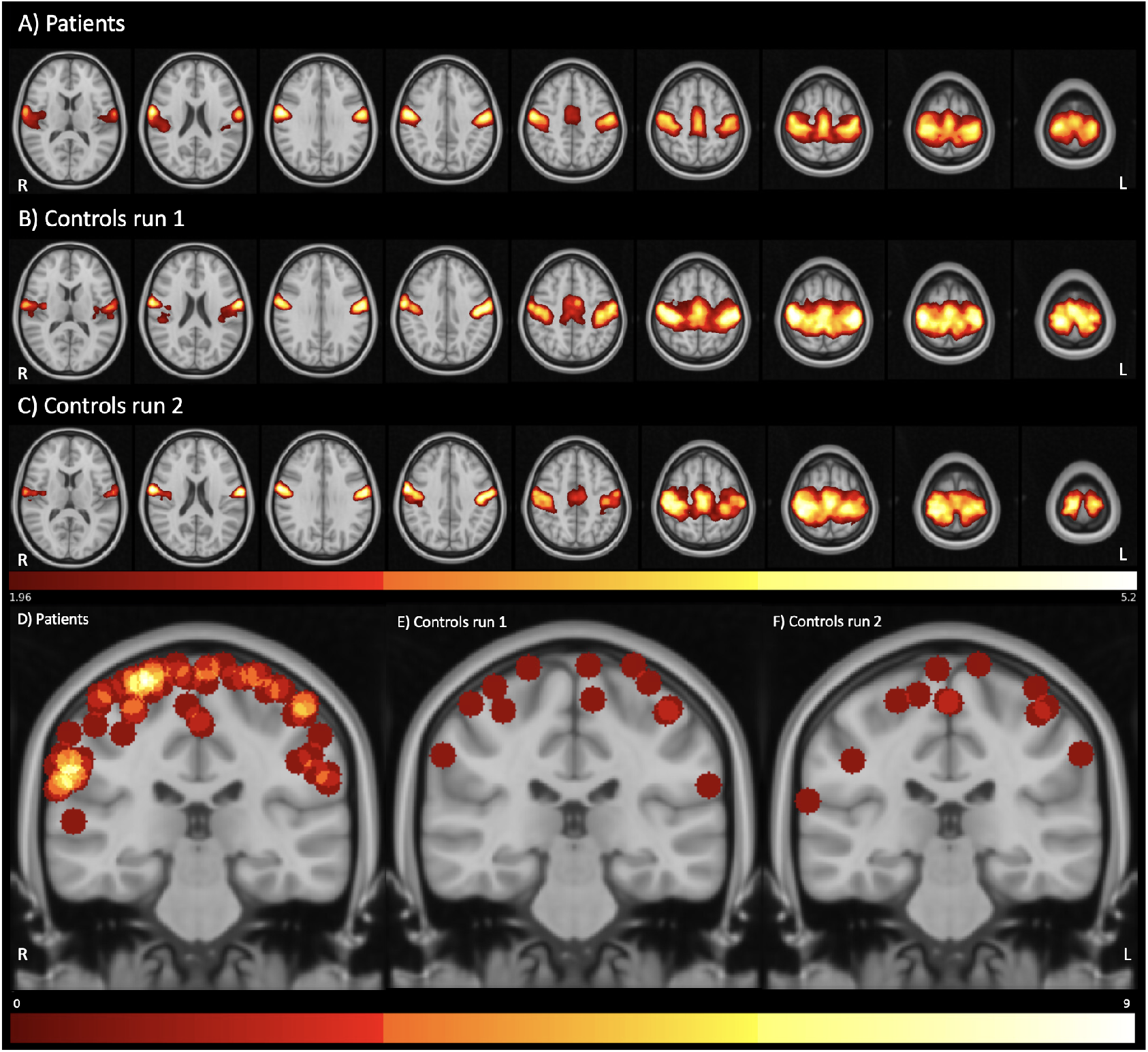
Motor mapping in patients and control. The group average of the whole brain independent component mapping (wIMM) in (A) 64 patients, (B) 12 controls first run, and (C) 12 controls second run represented as standardized z-value maps. Panels (D), (E), and (F) illustrate the distribution of individual maxima of the motor map for control and patients groups, respectively. All maxima were projected to the representative coronal slice with Y= -28 in MNI space. The color intensity represents the number of participants with their maxima in that location.

### Comparison to task activation

Eleven controls and 32 patients had finger tapping motor task results available for comparison to wIMM results. The mean overlap between wIMM and motor task activation at a threshold of p=0.05 ranged from 60-77% across control and patient participants. In the control group, all motor maps captured the peak task activation at subject level, i.e. sensitivity of 100%. In the patient group, the sensitivity was 87.5% (28/32). The four wIMMs that were not sensitive to the task peak were found to have a lower wIMM mean Z value compared to sensitive wIMMs (t(df) = -5.73(6.78), p = 0.0007). Examples of concordance between wIMM and the task activation is illustrated in a control with a high sensitivity (Figure 3a) and low concordance in a patient with low hit rate and sensitivity to task activation (Figure 3b). In two instances, our method mapped regions in the motor regions despite the failure of the motor task, i.e. no clear activation cluster in the expected region (Supplementary figure 1). We compared the wIMM and task fMRI results in a single patient who had a subsequent subdural grid placement, with motor mapping through direct cortical stimulation (Figure 4). A majority (7/11 controls and 23/32 patients) showed bilateral activation in their motor task fMRI as well as their wIMM results, defined as LI of -0.2 to 0.2. There were no differences between task activation and wIMM LI in patients (t(df) = - 0.02(31), p = 0.98) nor in healthy controls (t(df) = -0.45(10), p = 0.66). The asymmetry in wIMM and task results (the sign of LI) showed a trend towards the opposite of their hemisphere of onset (t(df) = -1.79(16.64) p = 0.09, supplementary figure 2).

**Fig. 3.**
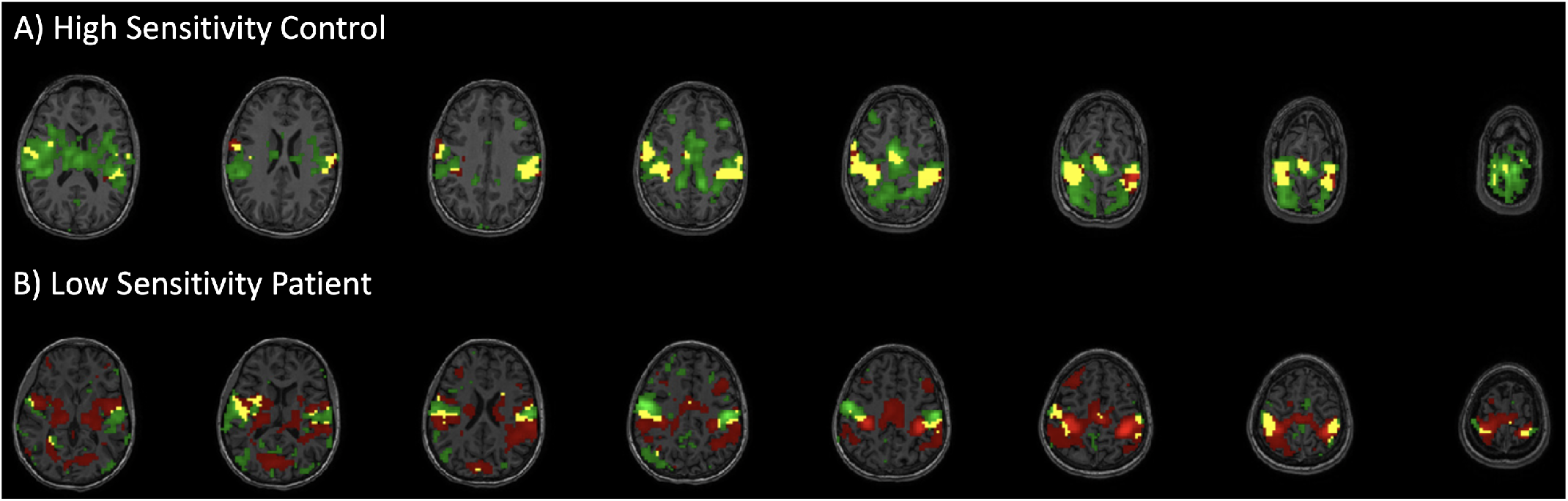
Concordance with task activation at subject level. Examples for high and low concordance between wIMM and task activation among participants. Panel (A) shows a control participant representing the high concordance seen in all controls (100%) and the majority (87.5%) of patients. Panel (B) shows a patient representing individuals with lower wIMM sensitivity for capturing the peak of the task activation. Although the maxima of the task activation and motor component map are spatially discrete, they both fall into the motor cortex regions. Green: whole brain ICA motor map (wIMM), Red: finger tapping motor task activation, and Yellow: overlap between maps corresponding to p<0.05.

**Fig. 4.**
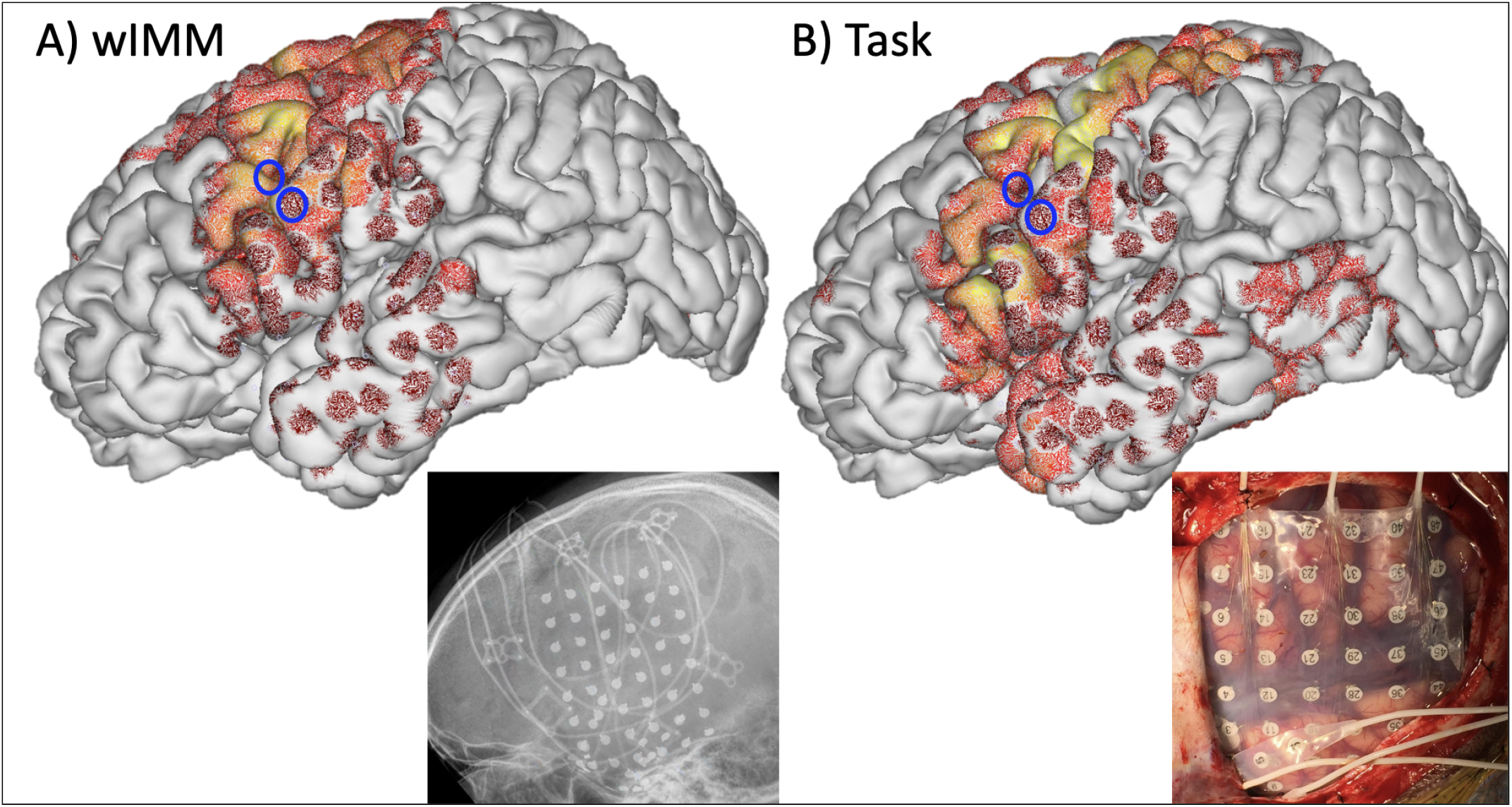
Case study. The results from whole-brain ICA-derived motor component map (wIMM, panel A) and finger tapping task activation (panel B) are compared for a left-handed child with refractory focal epilepsy and left temporal MTS. The 6×8 subdural grid electrodes are projected to the cortical surface and overlaid on the native pial surface mesh, and thumbnails show its position in lateral image and intra-operational picture. Blue circles indicate electrodes identified as motor cortex from clinical direct electrical stimulation motor mapping, which are in the expected precentral gyrus “hand knob” region. The wIMM results and task fMRI results are both thresholded at p<0.05. Visual comparison of the overlapping maps, and the proximity of the maxima for each map to the identified hand region shows high concordance at the individual level.

### Correlation with neuropsychological scores

Handedness scores (EHI) were available for all 64 patients, grooved pegboard data was available in 17 patients, and DKEFS motor trail-making data was available in 15 patients. The EHI score and wIMM peak value (r = 0.26, p = 0.04) were correlated. In contrast, EHI and task peak value were not correlated (r = 0.2, p = 0.27). The wIMM or task peak values did not correlate with DKEFS motor trail-making or Grooved Pegboard scores. Similarly, the laterality (LI) for wIMM and task activation did not correlate with any of those measures.

### Reproducibility

In healthy controls, we compared wIMM derived from two separate resting state runs for intra-subject reproducibility. The mean Dice coefficient across runs was 0.70 indicating high similarity, and the wIMM of 10/12 controls demonstrated a Dice coefficient of at least 0.60 between repeated runs (Supplementary figure 3). The mean wIMM z values in and across hemispheres did not differ between the two runs (t(df)= 1.06(11), p>0.3). Mean wIMM LI were not different between the two runs (−0.03 for first versus -0.07 for second run, t(df)= 1.01 (11), p = 0.33). This demonstrates stability of the method across resting state runs.

## Discussion

In this study, we optimized a whole-brain ICA-based method that uses rsfMRI to map clinically pertinent motor regions for children with epilepsy. Our results show concordance with finger tapping motor task fMRI, the current method of choice in most clinical settings. Overall, based on concordance with the task activation, we mapped the motor cortex in all 11 controls, and in 28 out of 32 epilepsy patients with motor task fMRI available for comparison at a threshold of z=1.96. Our results also show within-subject reproducibility of wIMM, and concordance with direct brain stimulation in one patient who underwent clinical direct electrical stimulation motor mapping (Figure 4). Using rsfMRI is highly relevant in surgical evaluation of children with epilepsy: accurate mapping is crucial for safe surgical planning. However, achieving good quality task activation can be challenging in young children, and comorbid cognitive problems may limit their ability to follow task instructions. Our rationale for choosing motor mapping as the proof of principle for our method lays in its robustness, which is well known since rsfMRI was first introduced(8, 25–27). Seed-based mapping of motor cortex adjacent to tumors by placing the seed on the healthy appearing contralateral precentral region can be comparable to direct cortical stimulation mapping (28). One study using ICA analysis of rsfMRI to delineate sensorimotor cortex in healthy adults and in four brain tumor patients showed the feasibility of the approach (16). Other studies have used ICA analysis for language mapping and compared it to task data, showing modest concordance (13, 15). We generated the IC and further processed them in native space. This is clinically relevant, allowing for juxtaposition of wIMM results to lesions or abnormalities in the patient’s brain, and coregistration to other imaging modalities during surgical work up. To account for the variability in the level of integration, we estimated the ICs with different preset number of components (TNC levels). Based on prior studies comparing DICI measure for component selection to other methods for automated template matching, we considered this method as both sensitive and specific. Huang et al. developed a toolbox that in part utilize DICI with templates developed for individual centers (23). Lu *et al*. (14) validated the DICI method for identifying language network in 10 adult controls and 7 patients with brain tumors. In this study, we adapted and optimized a similar approach for motor cortex mapping. Limiting the fMRI searching space to a priori region for component identification, also called a masked ICA approach (29), could be an efficient approach to exclude the potential influence of physiologic or other acquisition-related noise. In patients with lower network integration, our whole brain approach may not capture all the motor subcomponents as they may connect to other networks. Conversely, a masked approach may exclude the components that have shifted or extended to less common regions because of brain plasticity or developmental constraints, a plausible situation in childhood onset epilepsy. Whole brain approach also provides an opportunity to study those variability in network organization to correlate with clinical outcomes. Future studies may use masked ICA approaches as complementary to wIMM. We noted that the four patients with their peak task activation outside the estimated wIMM had the lowest wIMM mean z values compared to other patients (Supplementary Figure 4). This may indicate lower integration strength of the components of the motor network and lower overall integration of the patient’s functional networks at rest compared to others. Although we could not test this hypothesis, we postulate that this overall variability of the motor network integration may explain postsurgical deficits in motor function not predicted by conventional task fMRI. Conversely, there were instances where wIMM results mapped motor function while task fMRI was considered null (Supplementary figure 1). This may suggest that wIMM can identify extended regions involved with the motor network that may not be captures in task fMRI. We did not find any correlation between wIMM results and the available neuropsychology data related to motor function. Interestingly, the hemisphere of seizure onset in patients showed a small effect on the lateralization of the wIMM. This can be related to modulation of resting networks by epileptic networks in focal epilepsy, which by definition are asymmetric. A similar effect has been reported in studies on reorganization of language networks in children with epilepsy (30). Our study is likely underpowered for confirming those effects, but future studies including larger sample size may evaluate a possible effect of handedness and seizure focus side on functional motor map changes. Our study has a number of limitations. Factors such as post-surgical anatomy, physiologic noises, and role of medications or underlying conditions on rsfMRI signal variability are not fully controlled. This is a common pitfall in using real-world patient data, particularly pediatric population despite employing best practices in pre-processing and controlling for movement parameters. Using active resting state protocols (17), and advanced outlier identification methods may curtail those limitations in future studies. We had limited healthy control data, although we achieved fairly uniform results in concordance of wIMM and task results in controls. We did not have neuropsychological motor scores for all patients. Although hand region is often the most critical region for surgical decision-making, we did not look at other motor sub-regions, since we do not regularly collect task fMRI for leg or face. Analysis of other datasets with more granularity in motor tasks may evaluate the validity of our approach in the rest of the motor cortex.

## Conclusions

In summary, this study uses DICI measure to select the best ICs from whole brain rsfMRI data by matching to a clinically pertinent motor template. We observed a high concordance between our motor component maps and the task, demonstrated by high percentage of overlap, and high sensitivity of the motor component map for including the peak activation in both patients and controls. Our approach is data-driven and shows promising level of congruence with current standard of care in surgical work up for epilepsy surgery. Our results are an important step towards developing subject level clinical tools to map eloquent brain regions, including regions with more variability and complexity such as language in the presurgical evaluation of children with epilepsy using rsfMRI.

## Data Availability

All data produced in the present study are available upon reasonable request to the authors. A code repository is available for reproducibility.

https://github.com/tgholipour/ICAmotor.

## ACKNOWLEDGEMENTS

This study was supported by the NIH National Center for Advancing Translational Sciences (UL1TR001876/KL2TR001877 to TG), and the NICHD (Intellectual and Developmental Disabilities Research Center 1U54HD090257-01).

## Supplementary Materials section

- Supplementary Figure 1
- Supplementary Figure 2
- Supplementary Figure 3
- Supplementary Figure 4
- Supplementary Table

