## Supplemental for "Resting State fMRI for Motor Cortex Mapping in Children with Epilepsy"

#### Supplementary Figure 1

An example of a patient with a wIMM representing a motor network (A), while motor task fMRI does not capture any activation (B), also known as a null study.

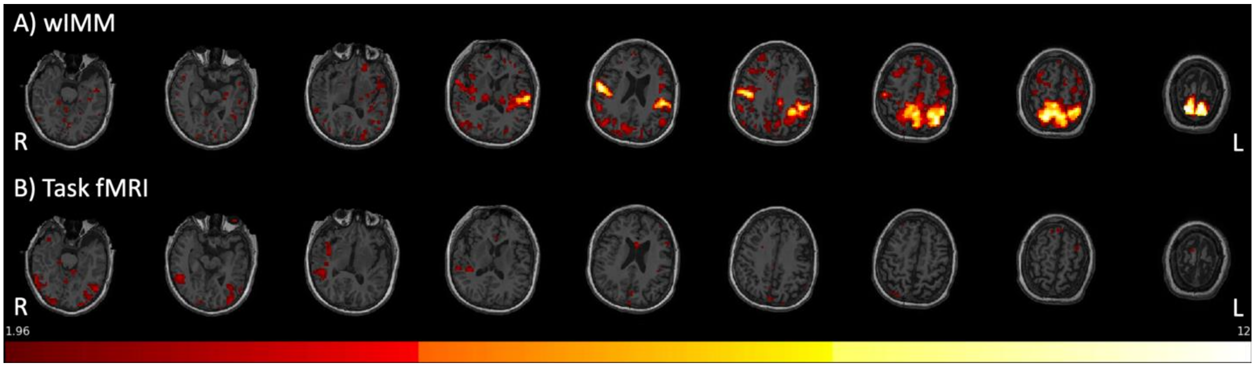

Supplementary Figure 2

The laterality index (LI) of the whole brain ICA motor map (wIMM) and motor task fMRI activation maps plotted based on the hemisphere of seizure focus. Despite a trend in wIMM LI showing shift towards the opposite hemisphere to the seizure focus, the differences did not reach significance. LI laterality designations are shaded (Left: LI>0.2 in green, Bilateral: |LI|<=0.2 in blue, Right: LI<-0.2 in red).

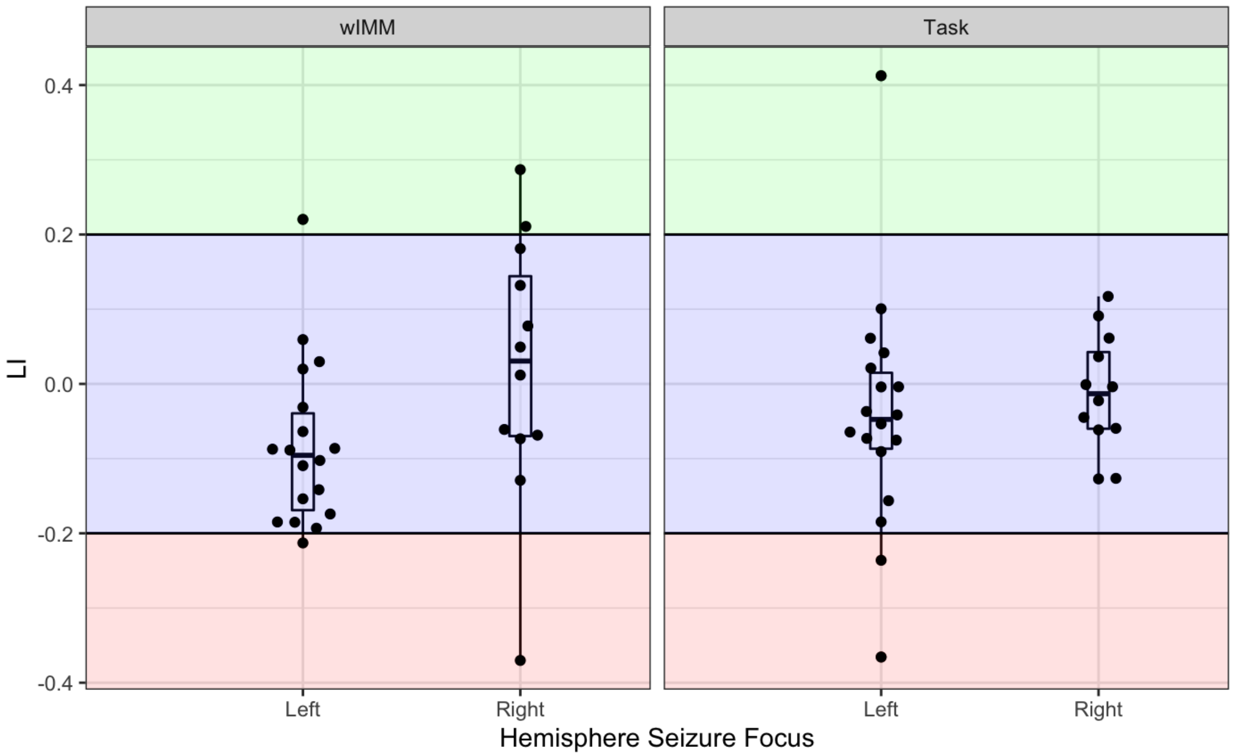

### Supplementary Figure 3

Reproducibility of the wIMM results Panel (A) shows the bilateral mean z value of the whole brain independent component motor map (wIMM) in two separate resting state runs for control group (N=12). Each line represents one subject. Panel (B) shows the distribution of dice coefficients between the wIMMs of repeated runs of controls.

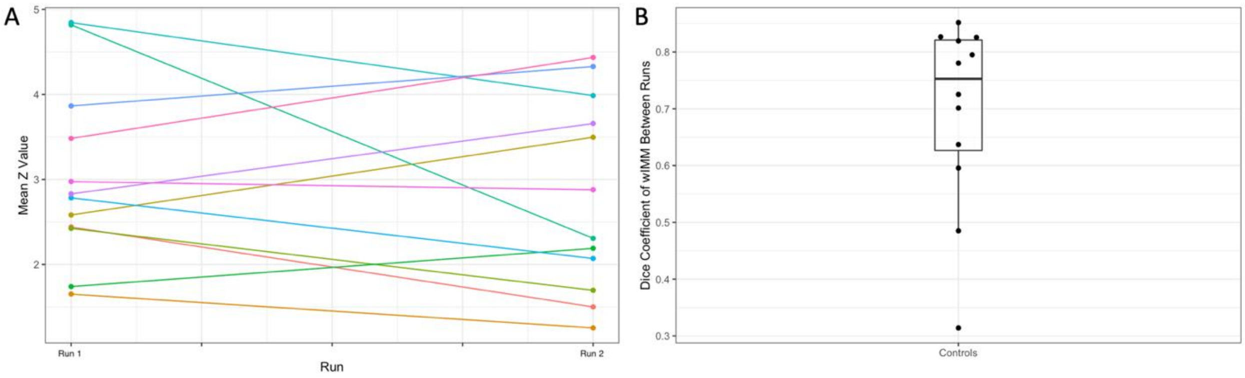

### Supplementary Figure 4

Four patients (A-D) with peak task activation outside of the wIMM. The estimated wIMM (green) for these four patients, did not include the peak of the task fMRI activation maps (hot) when thresholded at z = 1.96.

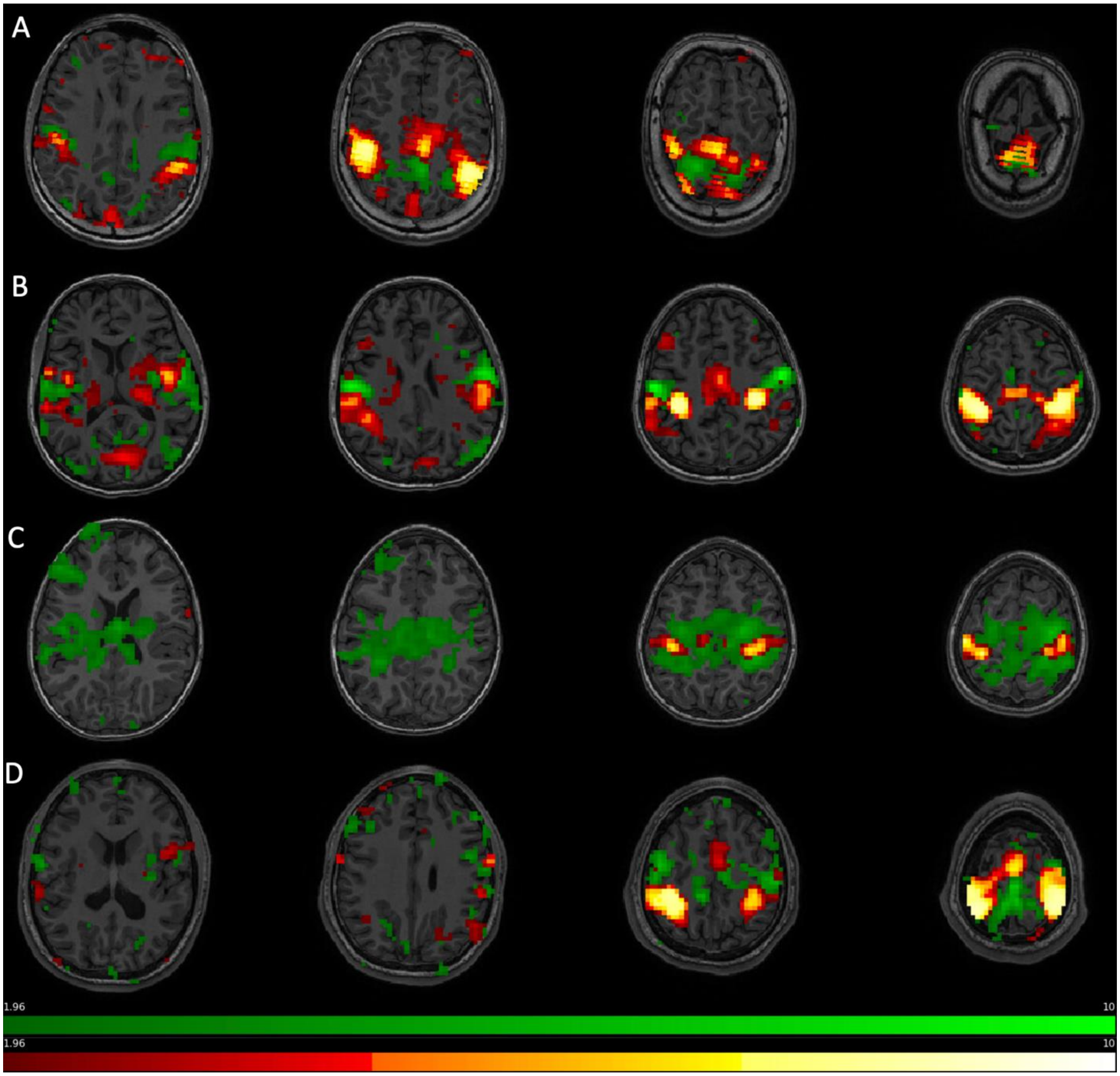

### Supplementary Table

Detailed demographic and clinical characteristics of participants. Ages are reported as age ranges in 5-year intervals.

| Subject # | Hemisphere of Onset | Handedness | Sex | Age at Scan | Lobe of Onset |
|---|---|---|---|---|---|
| 1 | Bilateral | Left | Male | 11-15 | Frontal, Temporal |
| 2 | Bilateral | Right | Female | 16-20 | Frontal, Temporal |
| 3 | Left | Ambidextrous | Female | 6-10 | Temporal |
| 4 | Left | Ambidextrous | Female | 11-15 | Temporal (Parietal, Frontal, Occipital) |
| 5 | Left | Ambidextrous | Female | >20 | Frontal, temporal |
| 6 | Left | Ambidextrous | Female | 16-20 | Frontal, Parietal, Temporal |
| 7 | Left | Ambidextrous | Male | 11-15 | Temporal |
| 8 | Left | Ambidextrous | Male | 16-20 | Temporal |
| 9 | Left | Ambidextrous | Male | 11-15 | Temporal |
| 10 | Left | Left | Female | 11-15 | Frontal |
| 11 | Left | Left | Female | >20 | Frontal, Temporal |
| 12 | Left | Left | Female | 6-10 | Central |
| 13 | Left | Left | Female | 11-15 | Temporal |
| 14 | Left | Left | Male | 15-20 | Temporal |
| 15 | Left | Left | Male | 11-15 | Frontal, Temporal |
| 16 | Left | Left | Male | 11-15 | Frontal |
| 17 | Left | Left | Male | 11-15 | Temporal |
| 18 | Left | Left | Male | 6-10 | Temporal |
| 19 | Left | Right | Female | 11-15 | Temporal |
| 20 | Left | Right | Female | 11-15 | Parietal |
| 21 | Left | Right | Female | 11-15 | Temporal, Parietal, Frontal |

| | | | | | |
|---|---|---|---|---|---|
| 22 | Left | Right | Female | 16-20 | Temporal, Frontal |
| 23 | Left | Right | Female | 11-15 | Frontal, Central |
| 24 | Left | Right | Female | 11-15 | Frontal |
| 25 | Left | Right | Female | 16-20 | Frontal, Temporal |
| 26 | Left | Right | Female | 11-15 | Frontal |
| 27 | Left | Right | Female | 11-15 | Frontal |
| 28 | Left | Right | Female | 11-15 | Temporal, Parietal |
| 29 | Left | Right | Male | 16-20 | Frontal |
| 30 | Left | Right | Male | 11-15 | Temporal |
| 31 | Left | Right | Male | 11-15 | Parietal |
| 32 | Left | Right | Male | >20 | Tempora, Parietal |
| 33 | Left | Right | Male | 11-15 | Frontal, Temporal |
| 34 | Left | Right | Male | 16-20 | Frontotemporal |
| 35 | Left | Right | Male | 11-15 | Frontal |
| 36 | Left | Right | Male | 11-15 | Tempora, Parietal |
| 37 | Left | Right | Male | 11-15 | Frontal (Parietal, Temporal) |
| 38 | Right | Ambidextrous | Female | 11-15 | Temporal |
| 39 | Right | Ambidextrous | Female | 6-10 | Frontal, Temporal |
| 40 | Right | Ambidextrous | Male | 11-15 | Temporal, Parietal |
| 41 | Right | Ambidextrous | Male | 11-15 | Parietal |
| 42 | Right | Left | Female | 6-10 | Temporal, Parietal |
| 43 | Right | Left | Male | 6-10 | Occipital |
| 44 | Right | Left | Male | 16-20 | Temporal, Frontal |
| 45 | Right | Left | Male | 16-20 | Temporal |
| 46 | Right | Right | Female | 16-20 | Frontal |
| 47 | Right | Right | Female | 11-15 | Frontotemporal |

| | | | | | |
|---|---|---|---|---|---|
| 48 | Right | Right | Female | 11-15 | Frontal |
| 49 | Right | Right | Female | 16-20 | Frontal |
| 50 | Right | Right | Female | 6-10 | Frontal, Temporal |
| 51 | Right | Right | Female | 11-15 | Temporal |
| 52 | Right | Right | Female | 16-20 | Frontotemporal |
| 53 | Right | Right | Female | 16-20 | Frontal, Temporal |
| 54 | Right | Right | Female | 11-15 | Parietal |
| 55 | Right | Right | Female | 6-10 | Parietal |
| 56 | Right | Right | Female | 11-15 | Temporal |
| 57 | Right | Right | Male | 6-10 | Frontal, Temporal |
| 58 | Right | Right | Male | 11-15 | Temporal |
| 59 | Right | Right | Male | 0-5 | Temporal |
| 60 | Right | Right | Male | 16-20 | Temporal |
| 61 | Right | Right | Male | 11-15 | Frontal |
| 62 | Right | Right | Male | >20 | Temporal |
| 63 | Right | Right | Male | 11-15 | Temporal |
| 64 | Right | Right | Male | 6-10 | Frontal |
